# Adenovirus and RNA-based COVID-19 vaccines: perceptions and acceptance among healthcare workers

**DOI:** 10.1101/2020.12.22.20248657

**Authors:** Mohamad-Hani Temsah, Mazin Barry, Fadi Aljamaan, Abdullah Alhuzaimi, Ayman Al-Eyadhy, Basema Saddik, Abdulkarim Alrabiaah, Fahad Alsohime, Ali Alhaboob, Khalid Alhasan, Ali Alaraj, Rabih Halwani, Nurah Alamro, Fatimah S Al-Shahrani, Amr Jamal, Sarah Alsubaie, Ziad A Memish, Jaffar A Al-Tawfiq

**Affiliations:** Pediatric Department, College of Medicine, King Saud University, Riyadh, Saudi Arabia; Division of Infectious Diseases, Department of Internal Medicine, College of Medicine, King Saud University and King Saud University Medical City, Riyadh, Saudi Arabia; College of Medicine, King Saud University, Riyadh, Saudi Arabia; Critical Care Department, College of Medicine, King Saud University, Riyadh, Saudi Arabia; Dr Sulaiman Al Habib Medical Group, Riyadh, Saudi Arabia; Division of Pediatric Cardiology, Cardiac Science Department, College of Medicine, King Saud University, Riyadh, Saudi Arabia; College of Medicine, University of Sharjah, Sharjah, UAE; Department of Medicine, College of Medicine, Qassim University, Qassim, Saudi Arabia; Department of Family and Community Medicine, King Saud University Medical City, Riyadh, Saudi Arabia; Evidence-Based Health Care & Knowledge Translation Research Chair, King Saud University, Riyadh, Saudi Arabia; Director Research and Innovation Centre, King Saud Medical City, Ministry of Health & College of Medicine, Alfaisal University, Riyadh, Kingdom of Saudi Arabia; Hubert Department of Global Health, Rollins School of Public Health, Emory University, Atlanta, GA, USA; Specialty Internal Medicine and Quality Department, Johns Hopkins Aramco Healthcare, Dhahran, Saudi Arabia; Infectious disease division, Department of Medicine, Indiana University School of Medicine, IN, USA; Infectious Disease Division, Department of Medicine, Johns Hopkins University School of Medicine, Baltimore, MD, USA

**Keywords:** COVID-19, vaccines, information source, anxiety, healthcare workers

## Abstract

**Objectives:** The aim of this study was to compare the perception, confidence, hesitancy, and acceptance rate of various COVID-19 vaccine types among healthcare workers (HCWs) in Saudi Arabia, a nation with MERS-CoV experience.

**Design:** National cross-sectional, pilot-validated questionnaire.

**Setting:** Online, self-administered questionnaire among HCWs.

**Participants:** A total of 2,007 HCWs working in the Kingdom of Saudi Arabia participated; 75.3% completed the survey and were included in the analysis.

**Intervention:** Data were collected through an online survey sent to HCWs during November 1-15, 2020. The main outcome measure was HCW acceptance of COVID-19 candidate vaccines. The associated factors of vaccination acceptance were identified through a logistic regression analysis and via measurement of the level of anxiety, using the generalized anxiety disorder 7 (GAD7) scale.

**Results:** Among the 1512 HCWs who were included, 62.4% were women, 70.3% were between 21 and 40 years of age, and the majority (62.2%) were from tertiary hospitals. In addition, 59.5% reported knowing about at least one vaccine; 24.4% of the participants were sure about their willingness to receive the ChAdOx1 nCoV-19 vaccine, and 20.9% were willing to receive the RNA BNT162b2 vaccine. However, 18.3% reported that they would refuse to receive the Ad5-vectored vaccine, and 17.9% would refuse the Gam-COVID-Vac vaccine. Factors that influenced the differential readiness of HCWs included their perceptions of the vaccine’s efficiency in preventing the infection (33%), their personal preferences (29%), and the vaccine’s manufacturing country (28.6%).

**Conclusions:** Awareness by HCWs of the several COVID-19 candidate vaccines could improve their perceptions and acceptance of vaccination. Reliable sources on vaccine efficiency could improve vaccine uptake, so healthcare authorities should use reliable information to decrease vaccine hesitancy among frontline healthcare providers.

## INTRODUCTION

The COVID-19 pandemic has severely disrupted normal societal and economic activities worldwide and is expected to continue imposing strains and burdens on health systems in most countries. Globally, the COVID-19 pandemic remains out of control.[1] The existing measures to control COVID-19 are detrimental to the global economy[2] and result in significant impairment in physical and psychological well-being.[3] To keep COVID-19 under control requires an effective vaccine. Without COVID-19 vaccination, healthcare workers (HCWs) will likely be at risk of infection and are likely to serve as a reservoir inside health institutes, which would undermine efforts to end the pandemic. According to the World Health Organization (WHO), 56 and 166 candidate vaccines are in clinical and pre-clinical evaluation, respectively, as of December 17, 2020.[4] These include JNJ-78436735, an adenovirus vaccine (Ad26.COV2.S)[5, 6]; mRNA-1273, an mRNA vaccine[7]; AZD1222, an adenovirus vaccine (ChAdOx1 nCoV-19)[8]; BNT162b1, an mRNA vaccine[9]; NVX-CoV2373, a full-length recombinant SARS CoV-2 glycoprotein nanoparticle vaccine adjuvanted with Matrix M[10]; and Ad5-nCoV, an adenovirus vaccine.[11-14] The encouraging news is that several vaccines have been released; many are in phase III clinical trials and show promising effectiveness.[15] As some safe and efficacious vaccines became available, policymakers must ensure successful, large-scale uptake of COVID-19 vaccines to achieve community immunization. However, the success of COVID-19 vaccination programs will largely depend on people’s acceptance of the vaccine. A recent global survey suggested that nearly 30% of participants would hesitate to take a COVID-19 vaccine when it is available.[16] A systematic review on the acceptance of a COVID-19 vaccine, based on nationally representative surveys in 20 nations, indicates that the vaccine acceptance rate in most nations would not reach the 67% necessary for achieving population immunity.[17] Mathematic modelling suggested that, if the efficacy of a COVID-19 vaccine was 80%, at least 75% coverage would be needed to extinguish the ongoing pandemic.[18] Therefore, a timely understanding of community responses to the forthcoming COVID-19 vaccines is important for policymaking and service planning.

Extant literature has explored vaccine acceptance and identified a few demographic and psychosocial correlates, such as gender, age, trust in research, knowledge, and concerns about the novel vaccine, as well as people’s judgment and perceptions about the risk of COVID-19.[19-21] Risk of exposure is one of several essential issues that directly shape people’s assessments of their vulnerability and risk. Even when using personal protective equipment, healthcare providers and other essential workers experience high-risk exposures to COVID-19 and should be given priority in vaccine allocations. Several studies suggest that being an HCW or being involved in the care of patients with COVID-19 is positively associated with COVID-19 vaccine acceptance.[22-24]

The lessons learned from previous infectious disease pandemics and outbreaks, including SARS, H1N1, MERS-CoV, and Ebola outbreaks, demonstrate the important role that health information has on disease control and vaccine acceptance.[25] Source of health information can affect the manner and frequency of the utilization of such information. The degree to which the information source is trusted can have a remarkable impact on the acceptance of information.[26] If HCWs distrust the source, they will doubt the information about different COVID-19 vaccines, and this doubt will in turn shape the attitudes, perceptions, and potential actions they take toward various COVID-19 vaccines.

The Kingdom of Saudi Arabia (KSA) is one of the top 30 countries with the highest reported COVID-19 cases: The KSA had 360,690 laboratory-confirmed cases and 6101 deaths as of December 19, 2020.[27] Acceptance of a potential COVID-19 vaccine assessed among HCWs in the KSA in a survey of 2007 participants showed an acceptance rate of 70%,[28] which is slightly higher than the acceptance rate found in a public survey among 992 participants from the general population (acceptance rate of 65%).[29] Perception of, confidence in, and hesitancy about various COVID-19 vaccines in the context of emerging viral infections and pandemics and with regard to manufacturing companies and different sources of information are principal factors in assessing vaccine acceptance. To the best of our knowledge, no published surveys specifically target and compare HCW perception, confidence, and hesitancy toward different types of COVID-19 candidate vaccines. Our previous research showed that most (70%) HCWs are willing to receive COVID-19 vaccines once they are available,[28] so we aimed in this study to compare the perception, confidence, hesitancy, and acceptance rates of various COVID-19 vaccine types among HCWs.

## METHODS

### Data collection

This study was a national cross-sectional survey among HCWs in Saudi Arabia during the COVID-19 pandemic. Data were collected during November 1–14, 2020. At the time of data collection, at least seven COVID-19 vaccine candidates had been reported in the scientific literature. HCWs were screened for their awareness of any of the seven published vaccines.[28] Participants were invited using a convenience sampling technique. We used several social media platforms and email lists to recruit participants. The survey was a pilot-validated, self-administered questionnaire that was sent to HCWs online through SurveyMonkey^©^, a platform that allows researchers to deploy and analyze surveys via the internet.[30] The questionnaire was adapted from our previously published study,[31] with modifications and additions related to the potential COVID-19 vaccine.

The questions included the demographic characteristics of respondents (job category, age, sex, years of clinical experience, and work area), and any previous exposure to Middle East respiratory syndrome coronavirus (MERS-CoV) or to patients with COVID-19 (either suspected or confirmed). We assessed the following outcomes related to the seven COVID-19 vaccine candidates that had been reported in the scientific literature: knowledge, perceived awareness, and readiness to receive each type of COVID-19 vaccine candidate. In addition, we assessed factors affecting respondents’ readiness to receive various COVID-19 vaccine candidates and the HCW’s sources of information about COVID-19 vaccines.

Before participation, the purpose of the study was explained in English at the beginning of the online survey. The respondent was given the opportunity to ask questions via a dedicated email address. The institutional review board at the College of Medicine and King Saud University Medical City approved the study (approval #20/0065/IRB). A waiver for signed consent was obtained because the survey presented no more than a minimal risk to participants and involved no procedures for which written consent is usually required outside the study context. To maximize confidentiality, personal identifiers were not required.

HCWs were screened for their awareness of any of the seven published vaccines. Notably, Pfizer announced during the study that the efficacy of their vaccine in the first interim analysis was more than 90%.[32]

### Statistical analysis

Descriptive statistics approaches, with means and standard deviations, were applied to continuous variables, and percentages were used for dichotomous variables. The two-sample *t* test was used to evaluate continuous scores, and the *Z* test was used to compare proportions.

A multivariable logistic regression model was used to explore associations between the outcome variable of HCW knowledge about the available COVID-19 vaccine candidates and HCW demographic, belief toward vaccine candidates, and level of anxiety. The association between predictors and the outcome was expressed as the odds ratio and 95% confidence interval. SPSS (version 21; IBM Corp) was used for the data analysis, Excel (Microsoft) was used for creating figures and depictions, and statistical significance was set at p=0.050. [33]

## RESULTS

A total of 2079 HCWs were invited to participate in the study; 2007 (96.5%) agreed to participate, and 1512 participants (75.3%) were included in the analysis. The participants’ sociodemographic characteristics are shown in Table 1.

**Table 1:** Respondents’ Sociodemographic and Professional Characteristics (N=1512)

| Characteristic | No. | % |
| --- | --- | --- |
| Sex |  |  |
| Male | 568 | 37.6 |
| Female | 944 | 62.4 |
| Age (years), mean (SD) |  | 37.28 (8.99) |
| 21-30 | 385 | 25.5 |
| 31-40 | 677 | 44.8 |
| 41-50 | 298 | 19.7 |
| ≥50 | 152 | 10.1 |
| Marital status |  |  |
| Single | 435 | 28.8 |
| Married, living with family | 715 | 47.3 |
| Married, living alone | 322 | 21.3 |
| Widowed or divorced | 40 | 2.6 |
| Any chronic illness |  |  |
| No | 1152 | 76.2 |
| Yes | 360 | 23.8 |
| Clinical role |  |  |
| Physician | 637 | 42.1 |
| Nurse | 757 | 50.1 |
| Other healthcare provider | 118 | 7.8 |
| Working area |  |  |
| Intensive care unit: adults and pediatrics | 216, 115 | 14.3, 7.6 |
| Emergency department | 152 | 10.1 |
| General ward | 406 | 26.9 |
| Isolation ward | 57 | 3.8 |
| Outpatient area | 319 | 21.1 |
| Other specialized unit: dialysis, lab, pharmacy, radiology | 206 | 13.6 |
| Hospital administrative | 41 | 2.7 |
| Hospital category |  |  |
| Private | 350 | 23.1 |
| Governmental | 712 | 47.1 |
| University hospital | 450 | 29.8 |
| Hospital level of care |  |  |
| Primary healthcare center | 210 | 13.9 |
| Secondary-care hospital | 361 | 23.9 |
| Tertiary hospital | 941 | 62.2 |

SD: Standard Deviation

Women comprised the majority of the population (62.4%), most participants (70.3%) were between 21 and 40 years of age, 68.6% were married—though only 47.3% were living with their families—and 76.2% reported not having any chronic illnesses. Respondents’ working areas were distributed almost evenly across different sectors of health institutions, but the majority were from the public/governmental (47.1%) sectors and tertiary institutions (62.2%). In terms of awareness of potential vaccine candidates reported in the literature, the majority (59.5%) reported knowing about at least one vaccine.

The ChAdOx1 nCoV-19 vaccine was the vaccine recognized the most by HCWs (39.3%), followed by the Gam-COVID-Vac vaccine (31.9%) and the RNA BNT162b2 vaccine (30.8%). The least well-known vaccine among HCWs was the mRNA-1273 vaccine (19.9%; Table 2).

**Table 2:** Perceived Awareness of and Readiness to Receive Various COVID-19 Vaccine Candidates by Healthcare Workers

|  |  |  |  |  |
| --- | --- | --- | --- | --- |
| Vaccine Candidate | No. (%) Knows About Vaccine* | No. (%) Ready to Take Vaccine |  |  |
|  |  | Never | Maybe | Sure |
| AstraZeneca (Oxford University: British/Swedish) non-replicating viral vector (chimpanzee adenovirus vectored vaccine (ChAdOx1 nCoV-19) | 594 (39.3) | 159 (10.5) | 984 (65.1) | 369 (24.4) |
| Gamaleya (Russia)-Sputnik V non-replicating viral vector adenovirus (Gam-COVID-Vac) | 482 (31.9) | 271 (17.9) | 1100 (72.8) | 141 (9.3) |
| Pfizer RNA (BNT162b2; USA): nucleoside-modified messenger RNA (modRNA) | 466 (30.8) | 154 (10.2) | 1042 (68.9) | 316 (20.9) |
| Johnson and Johnson (USA; adenovirus type 26 vector; Ad26.COV2-S) | 422 (27.9) | 154 (10.2) | 1108 (73.3) | 250 (16.5) |
| CanSino (China; adenovirus type 5; Ad5-vectored) | 397 (26.3) | 277 (18.3) | 1103 (72.3) | 132 (8.7) |
| Novavax (USA) protein subunit (full-length recombinant SARS CoV-2 glycoprotein nanoparticle vaccine adjuvanted with Matrix M; NVX-CoV2373) | 364 (24.1) | 166 (11) | 1139 (75.3) | 208 (13.8) |
| Moderna RNA (USA; mRNA-1273) | 301 (19.9) | 170 (11.2) | 1142 (75.5) | 200 (13.2) |

\*Percentage expressed of total sample (N=1512 healthcare workers).

HCWs were asked to indicate their readiness to receive each type of COVID-19 vaccine with response categories of never, maybe, or sure (i.e., willing to receive). The vaccine that most HCWs reported they were willing to receive was the AstraZeneca ChAdOx1 nCoV-19 (24.4%), followed by the Pfizer RNA BNT162b2 (20.9%) vaccine. Conversely, HCWs reported that they were most likely to refuse receipt of the CanSino Ad5-vectored (18.3%) and Gamaleya Gam-COVID-Vac (17.9%) vaccines. The respondents reported maybe most often for any vaccine candidate, with maybe responses ranging from 65.1% for the AstraZeneca vaccine to 75.5% for the Moderna mRNA vaccine (Table 2).

In determining the factors that influenced differential readiness of HCWs to receive the vaccine candidates that had been reported in scientific literature, a multiple-response dichotomies analysis showed that respondents’ perceptions of the vaccine candidate as more efficient in preventing infection was the most influencing factor (33%) in their decisions, followed by their personal preferences (29%) and the vaccine’s manufacturing country (28.6%). The least influential factors were media and social media coverage (12.3%) and trustworthiness (4.2%; Table 3).

**Table 3:** Factors Affecting Respondents’ Readiness to Receive COVID-19 Vaccine Candidates (N=1512)

|  |  |  |
| --- | --- | --- |
|  | No. | % |
| This COVID vaccine(s) seems more efficient in preventing the | 499 | 33 |
| infection. |  |  |
| Personal preference | 439 | 29 |
| Manufacturing country | 433 | 28.6 |
| Possibly fewer adverse effects from this vaccine | 417 | 27.6 |
| Vaccine availability | 394 | 26.1 |
| Company's reputation | 395 | 26.1 |
| Media coverage | 186 | 12.3 |
| Trustworthiness | 64 | 4.2 |

The HCW’s sources of information about COVID-19 vaccines are shown in Table 4. The WHO website was the most utilized source for information (51.1%), followed by social media networks (48.3%), the Saudi Ministry of Health (MOH) website (43.8%), and official press releases (38.3%). The Centers for Disease Control and Prevention website was utilized by only one third of participants (Table 4).

**Table 4:** Respondents’ Sources of Information About COVID-19 Vaccine Types (N=1512)

|  |  |  |
| --- | --- | --- |
|  | No. | % |
| WHO website | 762 | 51.1 |
| Social networks (e.g., YouTube, Facebook, Twitter, WhatsApp) | 719 | 48.3 |
| MOH website | 652 | 43.8 |
| Official statements or press releases from MOH (e.g., through SMS or newspapers) | 570 | 38.3 |
| Hospital announcements (e.g., roll-ups or newsletters) | 543 | 36.4 |
| Other internet resources | 537 | 36 |
| CDC website | 501 | 33.6 |

WHO: World Health Organization, MOH: Ministry of Health, SMS: Short Message Service, CDC: Centers for Disease Control and Prevention

A substantial number of HCWs in this study (n=612, 40.5%) reported unawareness of some vaccine candidates reported in scientific literature as of the time of the study. Therefore, as a secondary analysis, the generalized linear multivariate gamma regression analysis was used to explain the predictors of how likely the surveyed HCWs were to be aware of the different scientifically reported vaccine candidates. These results are presented in Table 5 and show that women knew significantly less than men about the different vaccine candidates (p=0.016). Older age correlated significantly and positively with more knowledge (p=0.027). Also, physicians knew significantly more about vaccine candidates than other HCWs did (p=0.001), and the HCWs from primary and secondary health centers knew of significantly fewer COVID-19 vaccine candidates than did HCWs from tertiary medical centers (p=0.002 for primary, p=0.02 for secondary). The participant’s belief in the ability of COVID-19 vaccines to stop the pandemic predicted significantly higher knowledge of the available vaccine candidates (p=0.009). HCWs who did not interact with COVID-19–infected family members knew significantly less about the available vaccine candidates (p=0.018). Other specific worry/anxiety levels and beliefs were assessed, as reported in Table 5

**Table 5:** Generalized Linear Modelling Analysis of the Healthcare Workers’ Knowledge of the Available COVID-19 Vaccine Candidates

| Parameter | Exponentiated ($\beta$) Coefficient | 95% CI for Exponentiated ($\beta$) | | p-value |
| --- | --- | --- | --- | --- |
|  |  | Lower | Upper |  |
| (Intercept) | 1.936 | 1.442 | 2.600 | <0.001 |
| Sex=female | 0.900 | 0.826 | 0.981 | 0.016 |
| Age (years) | 1.005 | 1.001 | 1.009 | 0.027 |
| Clinical role=physician | 1.267 | 1.101 | 1.458 | 0.001 |
| Clinical role=nurse and Midwife | 0.855 | 0.747 | 0.979 | 0.023 |
| Hospital setup type=primary | 0.847 | 0.763 | 0.940 | 0.002 |
| Hospital setup type=secondary | 0.904 | 0.830 | 0.984 | 0.020 |
| Hospital sector=private | 0.910 | 0.825 | 1.003 | 0.057 |
| Hospital sector=Governmental | 0.961 | 0.884 | 1.045 | 0.355 |
| Generalized anxiety, mean score | 1.002 | 0.995 | 1.010 | 0.565 |
| Worry level from getting COVID-19 viral infection, mean score | 0.961 | 0.920 | 1.005 | 0.080 |
| Worry level from transmitting COVID-19 viral infection to family, mean score | 1.029 | 0.991 | 1.069 | 0.133 |
| Believes the vaccine can stop the disease spread | 1.073 | 1.018 | 1.132 | 0.009 |
| Believes vaccination prevents COVID-19 complications | 1.046 | 0.994 | 1.100 | 0.087 |
| Does not interact with COVID-19–infected family members | 0.907 | 0.836 | 0.983 | 0.018 |
| NOTE: Dependent variable was the total number of vaccines the HCW knew about. The exponentiated ($\beta$) coefficient was interpreted as a rate. | | | | |

Our analysis (Fig 1) showed a significantly higher percentage rate of HCW readiness to receive any COVID-19 vaccine relative to the refusal rate after the Pfizer announcement compared to before it (χ^2^[1)]4.56, p=0.032). This result was similar to HCW readiness to receive the BNT162b2 vaccine (Fig 2).

**Figure 1:**
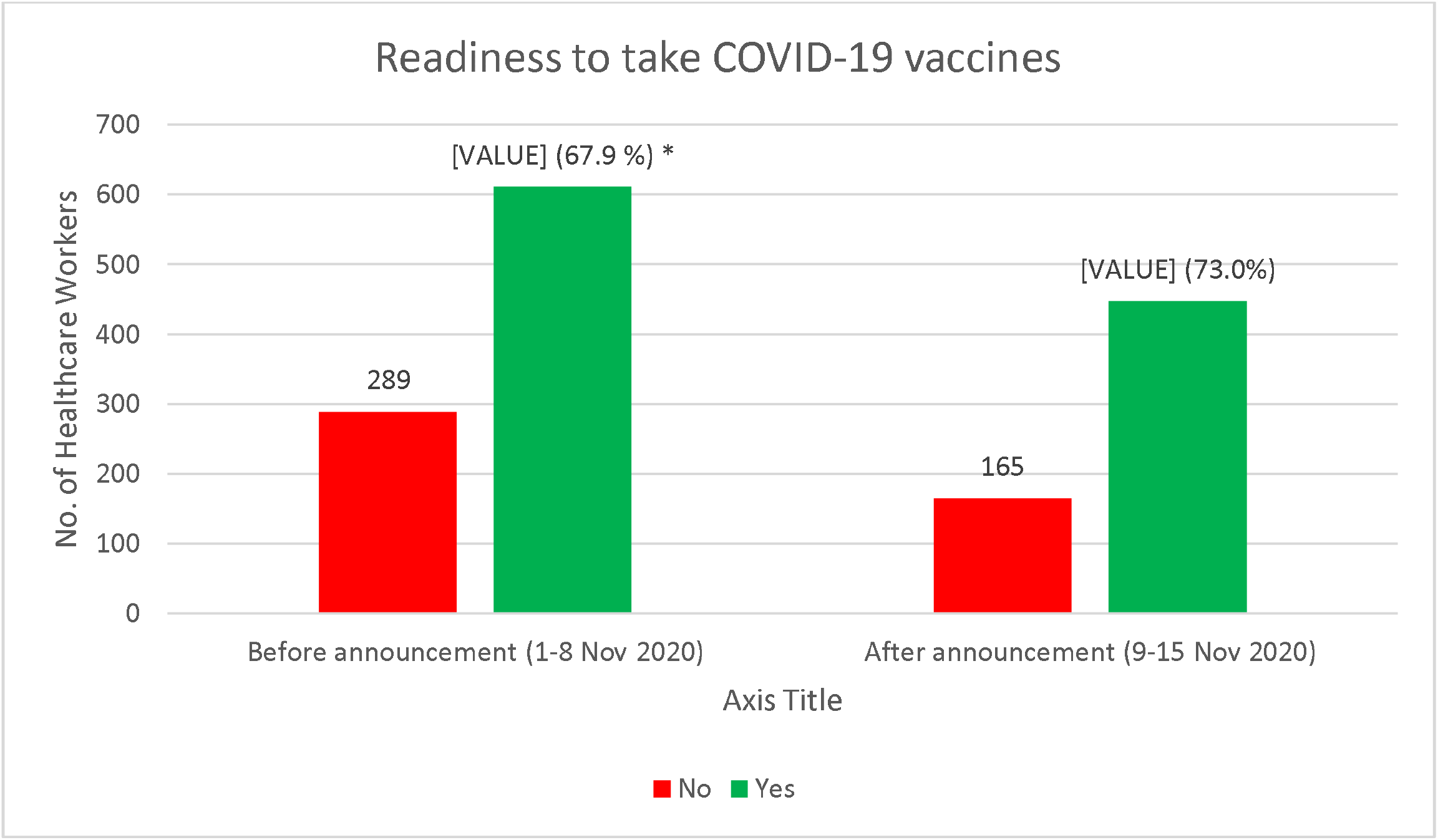
Readiness to take COVID-19 vaccines, as reported before and after the interim report of the efficacy rate of BNT162b2. *p=0.032.

**Figure 2:**
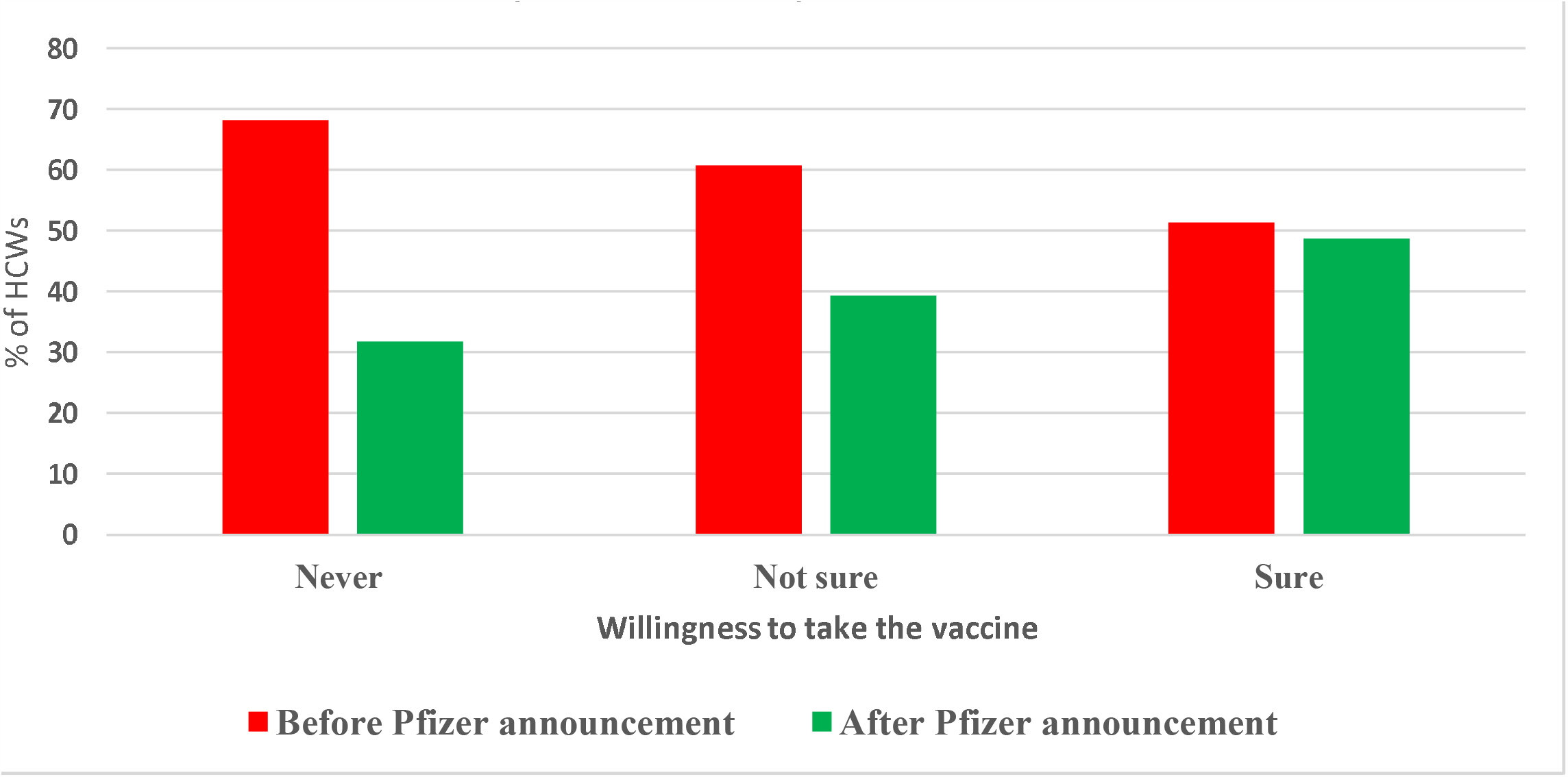
The percentage of healthcare workers (HCWs) willing to receive the BNT162b2 vaccine after its efficacy announcement. *p=0.001.

## DISCUSSION

Since the beginning of the pandemic, an unprecedented global effort to develop a vaccine has been underway; research and development of different technologies have been applied for different vaccine candidates. The effort resulted in several types of vaccine candidates developed with various technologies, including adenovirus and RNA-based vaccines, all of which are novel and have not been developed for wide clinical use in other infectious diseases. Gaining knowledge of such new vaccines, with the rapid evolution of the development process, may be challenging: only 40% of study participants were aware of the ChAdOx1 nCoV-19 vaccine,[5-9, 11, 34, 35] and only one third were aware of the BNT162b2, Gam-COVID-Vac, and Ad26.COV2-S vaccines. Only a quarter of participants knew about the remaining vaccines. To our knowledge, data about HCW knowledge of vaccine candidates has not been published elsewhere.

Acceptance about COVID-19 vaccines in general has been studied. In a global survey in 19 countries about the potential acceptance of a COVID-19 vaccine among the public, 71.5% reported they would very or somewhat likely agree to receive a vaccine; respondents from China gave the highest proportion of positive responses (631 [88.6%] of 712 respondents) and the lowest proportion of negative responses (five [0.7%0 of 712) when asked if they would take a proven, safe, and effective vaccine. Respondents from Poland reported the highest proportion of negative responses (182 [27.3%] of 666), whereas Russian respondents gave the lowest proportion of positive responses (373 [54.9%] of 680). Data are available about other diseases with multiple vaccine types as well. In a parental survey on acceptance of an intranasal, live, attenuated influenza vaccine, 81% preferred this version compared with the injectable inactivated influenza vaccine[36]; however no such acceptance rate has been evaluated among HCWs.[37]

It is interesting to note that, of all the HCW respondents asked about taking a COVID-19 vaccine, only 20% or 24% preferred to receive the AstraZeneca or the Pfizer vaccine, respectively. This low response to acceptance of any vaccine in development may indicate variability in the knowledge and understanding about the different vaccines. Vaccine knowledge is an area that needs more study to understand variables contributing to acceptance or rejection of each type of vaccine according to different development platforms used. This understanding would aid policymakers in the development of appropriate educational materials to boost confidence in various vaccine platforms.

Many factors affect the choice to receive vaccines. In this study, the top reason for choosing a vaccine was that the vaccine seems more effective at preventing infection (33%). A previous study found that 50% vaccine efficacy was associated with a 51% rate of acceptance.[38]

The manufacturing country was another reason given for accepting the vaccine (in 28.6% of respondents). This finding is similar to results from a US survey related to hypothetical vaccines. The surveyed individuals had lower acceptance of the vaccine if it originated from a country outside the United States.[38] Other contributing factors, such as fewer adverse effects, were also reported in this study.[38] Understanding these factors is important to build strategies for vaccine acceptance in any community. Strategies should address concerns, contributing factors, and misconceptions.[39] Trustworthiness was indicated by approximately 4% of the respondents as a factor in accepting a COVID-19 vaccine. It is important to note that trust is an important modifiable element of any successful vaccine campaign. Trustworthiness was strongly associated with acceptance of COVID-19,[40] and this factor was also related to acceptance of other vaccines, such as H1N1, SARS, and MERS vaccines.[25]

The most-reported sources of information for HCWs were the WHO website and social networks (as expected in a pandemic). Previously, Alsubaie et al.[41] reported results from the same HCWs’ population, which showed that hospital announcements and MOH official statements were more commonly sought for information about the MERS-CoV national outbreak. Seeking knowledge from reliable sources about the pandemic and vaccinations could significantly impact the HCWs’ perceptions of vaccine acceptance. Misinformation about the COVID-19 vaccine was associated with decreased vaccination acceptance among those who would otherwise definitely vaccinate.[42]

Interestingly, HCWs working in tertiary and academic centers were more knowledgeable about various vaccine candidates compared with HCWs working in primary and secondary centers. This result may be explained by more scientific activity and educational campaigns typically associated with teaching hospitals. This increased knowledge was especially common among physicians in our study, like other studies; in a cross-sectional survey conducted in Italy among HCWs to assess their knowledge, attitudes, and practices about vaccinations, physicians and those who had received information about vaccinations from scientific journals, educational activities, or professional associations were more likely to have adequate knowledge.[43] The knowledge differences identified between centers and types of providers highlight the importance of academic activities and keeping up-to-date with the scientific literature during the COVID-19 pandemic.

Remarkably, after the Pfizer and BioNTech announcement about the efficacy rate of BNT162b2, the HCWs in our study demonstrated significantly more willingness to undergo vaccination.[33] This change was despite simultaneous negative news on some COVID-19 vaccination trials, such as the halting of clinical studies with the CoronaVac vaccine by the Brazilian national sanitary regulator (Anvisa) due to a serious adverse event.[44] Vaccine acceptance is a multifactorial issue, but having positive COVID-19 vaccine trial results circulating in the news and social media for several days after the press release on the efficacy of BNT162b2 could improve the HCWs’ willingness to vaccinate.

## CONCLUSION

HCW awareness of the several COVID-19 candidate vaccines could improve perception and acceptance of vaccination. Reliable sources on vaccine efficiency could improve vaccine uptake, and healthcare authorities should utilize these sources to decrease vaccine hesitancy among frontline healthcare providers.

## Data Availability

Data is available upon reasonable request

## Acknowledgement

The authors are grateful to the Deanship of Scientific Research, King Saud University, for funding through the Vice Deanship of Scientific Research Chairs.

## Notes

### Competing Interest Statement

The authors have declared no competing interest.

### Author Declarations

The IRB at King Saud University, Riyadh, Saudi Arabia, approved the study (approval #20/0065/IRB)

